# Micro-heterogeneity of transmission shapes the submicroscopic malaria reservoir in coastal Tanzania

**DOI:** 10.1101/2023.09.06.23295089

**Authors:** Tyler Rapp, Kano Amagai, Cyrus Sinai, Christopher Basham, Mwajabu Loya, Sifa Ngasala, Hamza Said, Meredith S. Muller, Srijana B. Chhetri, Guozheng Yang, Ruthly François, Melic Odas, Derrick Mathias, Jonathan J. Juliano, Feng-Chang Lin, Billy Ngasala, Jessica T. Lin

## Abstract

**Background:** Asymptomatic malaria may be patent (visible by microscopy) and detectable by rapid malaria diagnostic tests (RDTs), or it may be submicroscopic and only detectable by polymerase chain reaction (PCR).

**Methods:** To characterize the submicroscopic reservoir in an area of declining malaria transmission, asymptomatic persons >5 years of age in Bagamoyo District, Tanzania, were screened using RDT, microscopy, and PCR. We investigated the size of the submicroscopic reservoir across villages, determined factors associated with submicroscopic parasitemia, and assessed the natural history of submicroscopic malaria over four weeks.

**Results:** Among 6,076 participants, *Plasmodium falciparum* prevalence by RDT, microscopy, and PCR was 9%, 9%, and 28%, respectively, with roughly two-thirds of PCR-positive individuals harboring submicroscopic infection. Adult status, female gender, dry season months, screened windows, and bednet use were associated with submicroscopic carriage. Among 15 villages encompassing 80% of participants, the proportion of submicroscopic carriers increased with decreasing village-level malaria prevalence. Over four weeks, 23% (61/266) of submicroscopic carriers became RDT-positive and were treated, with half exhibiting symptoms. This occurred more frequently in villages with higher malaria prevalence.

**Conclusions:** Micro-heterogeneity in transmission impacts the size of the submicroscopic reservoir and the likelihood of submicroscopic carriers developing patent malaria in coastal Tanzania.

## Introduction

Sub-Saharan Africa carries 95% of the world’s malaria burden (“Global messaging: World malaria report 2022,” n.d.). Despite substantial progress over the last two decades, persistent high burden areas continue to require resources and attention, while eliminating malaria from regions where transmission has declined has proven just as challenging. In particular, the asymptomatic reservoir that persists even after symptomatic malaria cases have declined poses a significant obstacle to malaria elimination. This asymptomatic malaria reservoir is dominated by low density infections not detectable by commonly used diagnostic tests (rapid diagnostic tests, or RDTs, and microscopy), often referred to as submicroscopic infections (Lin et al., 2014; Okell et al., 2012; Whittaker et al., 2021). While abundant studies have established the widespread prevalence of submicroscopic malaria in lower transmission settings, the factors that drive such prevalent asymptomatic carriage still remain elusive (Lindblade et al., 2013; Whittaker et al., 2021). Furthermore, studies have just begun to characterize the natural history of such infections (Barry et al., 2021; Sumner et al., 2021b), which is important for understanding their impact on human health and their role in ongoing transmission.

Some regions in East Africa have undergone an epidemiologic shift towards lower malaria endemicity over the last decade (Whittaker et al., 2021). Rural Bagamoyo district in coastal Tanzania represents one such area with previously moderate to high transmission that has experienced decreasing numbers of cases since the mid 2000s due to enhanced malaria control efforts. This epidemiologic transition is reflected in the highest incidence of symptomatic malaria cases occurring in older children and adolescents (10-19yo) who are exposed to malaria later in life vs. 0-9yo previously (Rutta et al., 2012). We conducted prospective cross-sectional malaria screening in Bagamoyo over three years as part of a parent study (Project TranSMIT) designed to investigate the asymptomatic infectious reservoir. Here we report the characteristics of the screening cohort and assess individual and ecological factors associated with submicroscopic parasite carriage. Since diagnosis of submicroscopic infection is usually limited to a slice in time, we sought to better understand the natural history of those with submicroscopic infection over four weeks of follow-up. We treated anyone who developed patent malaria or symptoms attributable to malaria during that time and otherwise deferred treatment to those that remained PCR-positive at the end of four weeks. The resulting comprehensive and nuanced portrait of the asymptomatic reservoir highlights significant micro-heterogeneity in a region of declining malaria prevalence.

## Methods

### Study site

The study was conducted in Bagamoyo District, Tanzania, a rural area on the eastern coast approximately 40 kilometers north of Dar es Salaam. This is an area of historically high malaria transmission that has undergone an epidemiologic transition to lower transmission over the last 15 years (Bhatt et al., 2015; Whittaker et al., 2021). Malaria transmission occurs throughout the year, with peaks typically during the long (March to June) and short (October to November) rainy seasons. Study data were collected as part of a large observational study called Project TranSMIT (Transmission of Submicroscopic Malaria in Tanzania) that prospectively screened and enrolled participants over three years for mosquito feeding studies and performed weekly follow-up over four weeks for a subset of participants with submicroscopic parasitemia (Markwalter et al., 2021; Tarimo et al., 2022).

### Participant recruitment and study procedures

We screened asymptomatic persons 6 years of age and older who denied antimalarial use within the previous 7 days. Participants who provided informed consent were screened at four primary schools in Bagamoyo district (Fukayosi, Mtakuja, Mkenge, and Mwavi) and two health centers (Yombo clinic and Fukayosi clinic), with the majority of participants recruited from Mwavi school and Yombo clinic. Both parental/guardian consent and child assent to participate was obtained for all children less than 18 years of age. Screened participants reported their age, gender, and village in which they reside. Approximately 200 μL of finger-prick blood was used to make thick and thin blood smears, perform a dual antigen HRP2 and pLDH RDT (rapid diagnostic test, SD Bioline), and create 2-3 dried blood spots (DBS) on Whatman 3MM filter paper for analysis by real-time polymerase chain reaction (PCR) targeting P. falciparum 18S rRNA as previously described (Markwalter et al., 2021; Tarimo et al., 2022). We previously determined the limit of detection (LOD) for the P. falciparum PCR test performed onsite as approximately 1-5 parasites/uL, which is more sensitive than both microscopy (approximately 50-500 parasites/uL) and RDT (approximately 100 parasites/uL), but not as sensitive as ultrasensitive PCR tests (Berzosa et al., 2018; Leski et al., 2020; Markwalter et al., 2021; Mfuh et al., 2019).

Individuals who were malaria positive by RDT or PCR were invited to enroll in the parent TranSMIT study for which they completed a further questionnaire covering housing and bednet use. Participants were excluded from enrollment if they were less than 6 years of age, reported fevers or chills within the previous 24 hours, were currently febrile, or had a serious illness. Those who screened RDT-positive were treated with artemether-lumefantrine according to Tanzanian national guidelines, either immediately or after enrollment procedures were completed on the same day. Enrolled participants who were RDT-negative/PCR-positive and not pregnant were further invited to complete longitudinal follow-up for 4 more weeks, including weekly follow-up for symptoms and repeat parasite testing at weeks 2 and 4 and upon report of any symptoms attributable to malaria (fevers, chills, headache, body aches, malaise, nausea/vomiting). These participants with asymptomatic submicroscopic parasitemia were provided with a thermometer to measure their temperature and were promptly treated if they developed malaria symptoms, became RDT or smear-positive, or at the end of 4 weeks if they remained parasite-positive by PCR. This study was approved by institutional review boards at the University of North Carolina, Tanzania National Institute for Medical Research, and Muhimbili University of Health and Allied Sciences.

### Data collection

Participant responses, study visit completion, and the results of parasite testing were entered into a REDCap database with verification by a second person. GPS coordinates of villages, local health facilities, and schools were obtained by the study team using an Android phone-based application (“Geo Tracker”), as well as through various existing geospatial data sources [5-8]. Rainfall data were obtained from the Tanzania Meteorological Agency (“Mwanzo,” n.d.).

### Statistical analyses

We defined submicroscopic parasitemia as low density infection detected by PCR that was not detected by RDT, reasoning that conventional HRP2-based RDTs generally detect parasite densities down to 100-200 parasites/μL, whereas the limit of detection of microscopy varies based on the quality of prepared smears, time devoted to diagnosis, and experience/expertise of readers (Berzosa et al., 2018; Das et al., 2017; Okell et al., 2012). Conversely, we use the term “patent malaria” to indicate PCR-positive infections that were also RDT-positive.

We report baseline characteristics for both screened and PCR-positive enrolled participants. Among those with PCR-positive infection, we compared the prevalence odds ratio of submicroscopic vs. patent malaria across key individual-level factors using chi-square, Mann-Whitney U, or Fisher’s exact tests as appropriate. The distribution of parasite densities was examined by season on the basis of rainfall, in which the dry season was defined as <33mm of rain per month and wet was defined as >33mm of rain per month (Gamoyo et al., 2015).

The proportion of asymptomatic PCR-positive infections that were submicroscopic (submicroscopic malaria ratio) was mapped at the village level alongside study area health facilities and schools using ArcGIS Pro 2.7 software. The relationship between malaria prevalence and the submicroscopic ratio was analyzed using linear regression models, with the number of participants and median age of participants sampled from each village, as well as geographic assignment to one of two catchment areas (Mwavi school or Yombo clinic) included as covariates.

The parasite status of participants with submicroscopic parasitemia were assessed per study week to demonstrate if participants became RDT-positive, continued to have submicroscopic infection, or became PCR-negative. Initial parasite densities and the trajectory of parasite densities among participants with these three outcomes were examined. Individual and village-level factors associated with development of RDT-positive patent malaria versus persistent carriage or resolution of parasitemia were analyzed using chi-square or Mann-Whitney tests as appropriate.

Analyses were performed using R 4.0.3 and GraphPad Prism 9. All hypothesis tests were two-sided at a significance level of 0.05 with no adjustment for multiple comparisons.

## Results

### Malaria prevalence and participant characteristics

From October 2018 to November 2021, 6,464 school-age children and adults in rural Bagamoyo district were screened for malaria using RDT, microscopy, and real-time PCR. Screening initially occurred during the rainy season months but was continuous for the latter half of the study except for a 7-month screening pause in 2020 due to the COVID-19 pandemic (Figure 1A). After excluding those who reported fever or other potential malaria symptoms in the previous 3 days (n = 250), those with suspected false positive RDT results (RDT-positive but PCR-negative, n = 95), and those who were previously already screened in a given season, 6,076 participants were included in our individual level analysis (Supplementary Figure 1).

**Figure 1.**
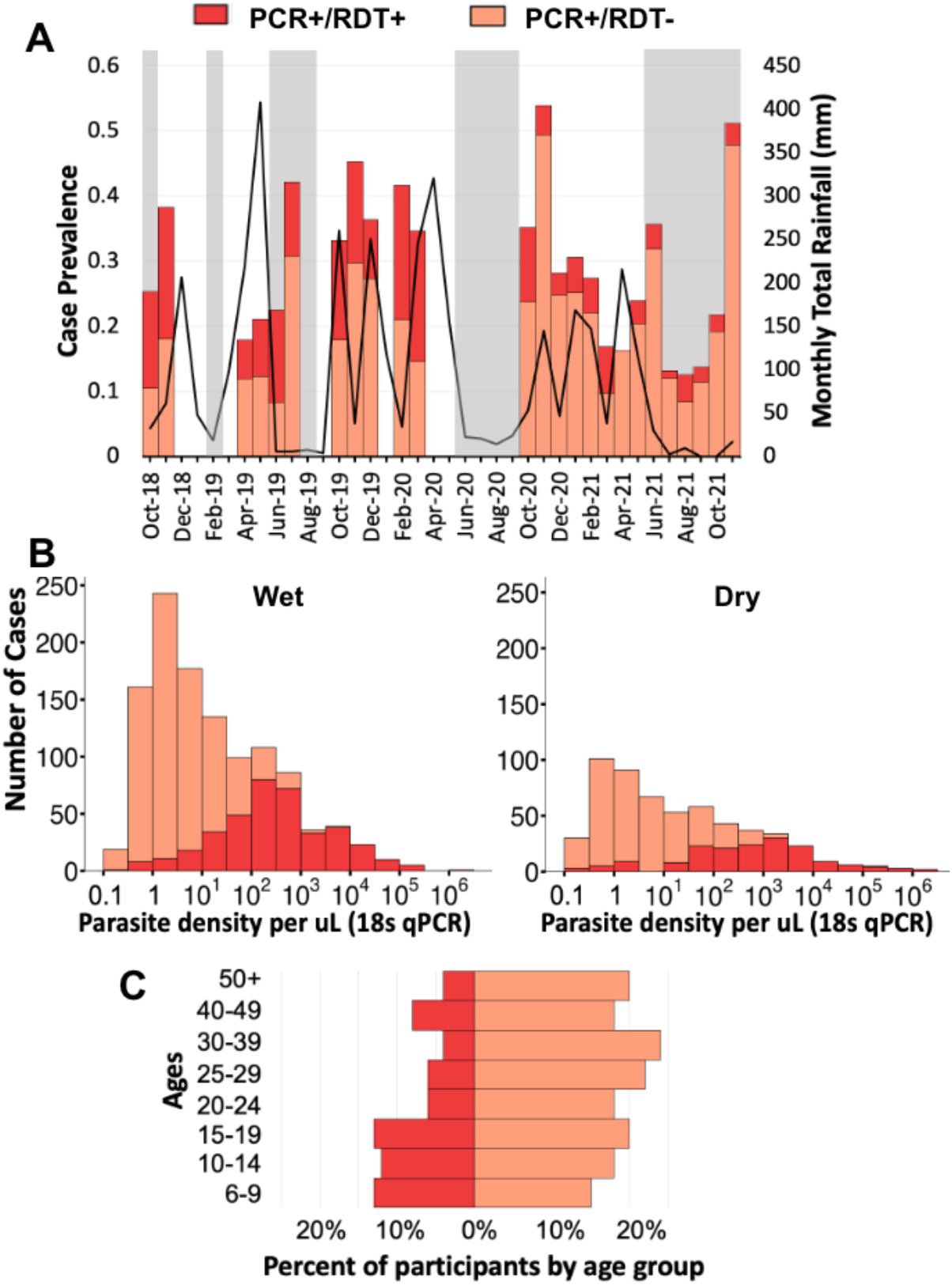
RDT-positive and submicroscopic malaria prevalence in relation to rainfall and age in Bagamoyo, Tanzania. Data are derived from screening 6,076 asymptomatic persons aged 6 and above between 2018-2021 using rapid diagnostic tests and 18S rRNA real-time PCR. Sampling was discontinuous. Dry season months based on lower rainfall (<33 mm) are indicated in gray in panel A. The distribution of parasite densities as determined by PCR in wet and dry season months is depicted in panel B. The proportion of participants who who tested positive by RDT vs PCR only for each age category is depicted in panel C.

Screened participants ranged in age from 6 to 100 years (median 19 years) (Supplementary Table 1). The majority (69%) were female, as women were more likely to accompany their children for screening or be recruited when bringing their children to clinic. Among this screening population, overall malaria prevalence by RDT, microscopy, and PCR was 9% (95% CI 8.4-9.8%), 9% (95% CI 7.9-9.3%), and 28% (95% CI 27-29%), respectively. Nineteen percent had submicroscopic infection, here defined as parasitemia detected only by PCR and not by RDT. Therefore, 68% (1,168/1,722), or two-thirds of asymptomatic infections, were submicroscopic. As expected, lower parasite densities (<100 parasites/μL), as determined by PCR, were largely subpatent and not detected by RDT (Figure 1B).

Gametocyte carriage identified by microscopy was relatively rare, occurring in 1.3% (78/6,076) of screened participants, 4.4% of those who screened malaria-positive by PCR, and 11% of those with RDT-positive patent infection. Further data were collected in a subset of those who screened malaria-positive and were enrolled in the parent TranSMIT study (n = 544) (Supplementary Table 1). The vast majority (89%) of these malaria-positive participants reported sleeping under a mosquito net the previous night. The majority lived in houses with sheet metal roofing (88%), built with bamboo and mud (68%) or cement (23%), with either open (43%) or screened windows (36%).

### Factors associated with submicroscopic parasitemia

Compared to participants who were RDT-positive, submicroscopic carriers were older (median age 21 vs. 13 years, *p* <0.001) and reported fewer malaria infections in the preceding year (one vs. two episodes, *p* = 0.002) (Table 1). Among younger school-age children, RDT-detectable patent malaria was common, but by young adulthood, submicroscopic infection prevailed (Figure 1C). Overall, adults had 2.3 times the odds of harboring submicroscopic malaria compared to school-age children 6-17 years of age. Interestingly, compared to malaria-positive males, malaria-positive females were more likely to harbor submicroscopic malaria (OR = 1.6 (95% CI 1.3-2.0)) (Table 1).

**Table 1.**
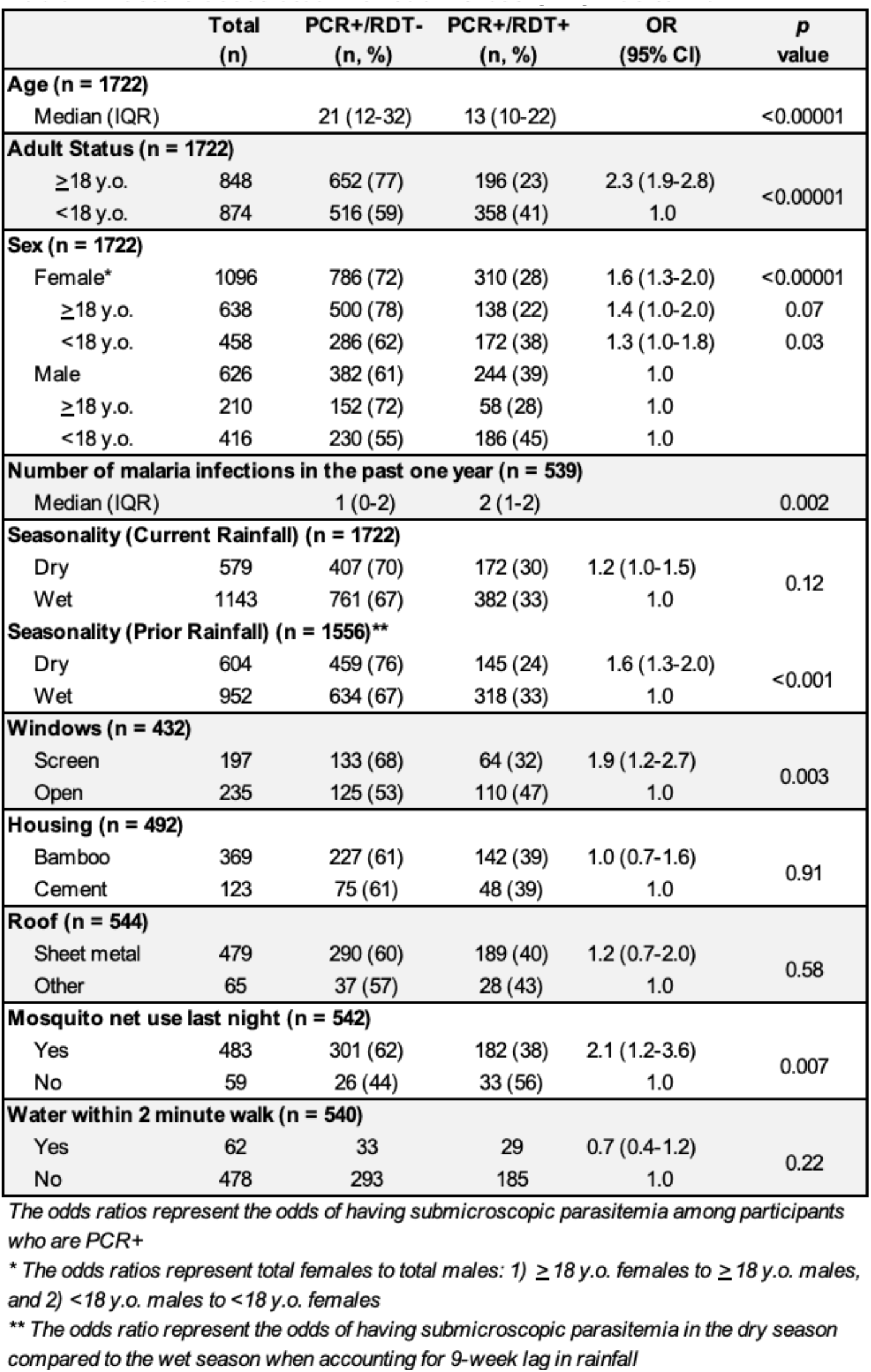
Factors associated with submicroscopic parasitemia.

In terms of ecological risk factors, malaria acquired after drier months and in the context of screened windows and bed net use was more likely to be submicroscopic. Seasonality, when divided into wet (>33mm rain) vs. dry (<33mm rain) months based on current rainfall, did not appear associated with the likelihood of patent malaria (Table 1). However, when prior rainfall was assessed, based on previous modeling showing a 9-week lag in the impact of rainfall on malaria case incidence (Krefis et al., 2011), dry months following periods of less rainfall (in the month falling 9 weeks prior) were associated with 1.6 (95% CI 1.3-2.0) times increased odds of submicroscopic malaria. Those living in dwellings with open windows were more likely to have patent malaria, whereas those with screened windows, and those who had used a mosquito net the previous night were more likely to have submicroscopic parasitemia (OR = 1.9 (95% CI 1.2-2.7), OR = 2.1 (95% CI 1.2-3.6), respectively).

The proportion of submicroscopic malaria was greater during the second half of screening (83% (626/750) from 2020-2021 vs. 56% (542/972) from 2018-2019), but differences in screening months and rainfall make this difference hard to interpret (Figure 1A). Stratified analysis of associated factors by study period shows that older age was more strongly associated with submicroscopic carriage in the first half of the study, whereas the association with current and prior rainfall differed in the two study periods (Supplementary Table 2).

### Size of the submicroscopic reservoir

We sought to determine if the size of the submicroscopic reservoir was related to transmission intensity, as indicated by local malaria prevalence at the village level. Study participants resided in an area spanning approximately 8,000 square kilometers across rural Bagamoyo district, with recruitment from two primary catchment areas about Mwavi and Yombo (Figure 2) (Japan International Cooperation Agency (JICA), Global Environment Department, 2005; Maina et al., 2019; Ministry of Health, The United Republic of Tanzania, 2021; OpenStreetMap contributors, Geofabrik GmbH, 2021; Tanzania Communications Regulatory Authority (TCRA), 2019). Among villages where participants reported residing, we selected the 15 villages with at least 75 screened participants that included both children and adults for analysis. This comprised 80% (4,852/6,076) of our total study sample.

**Figure 2.**
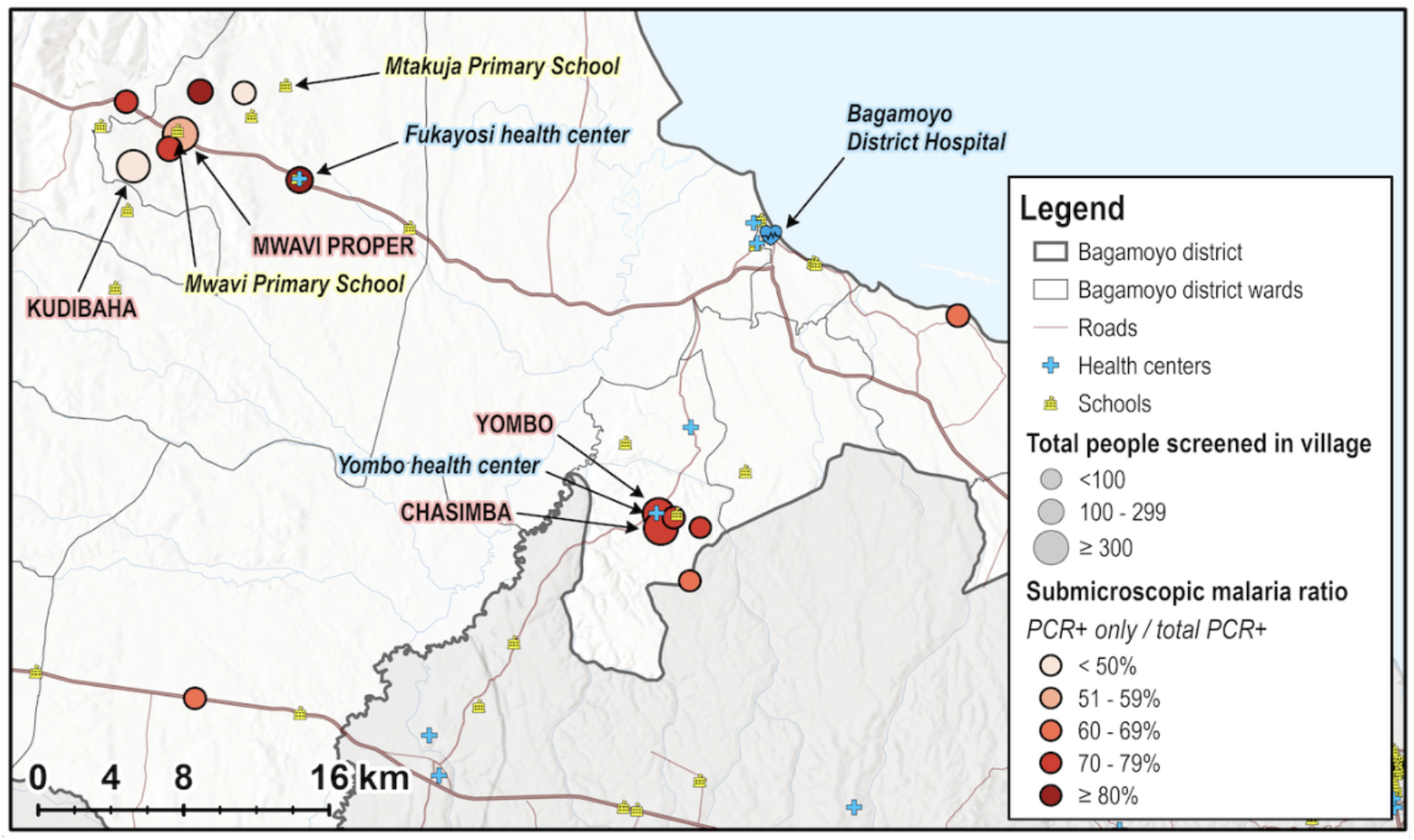
Village-level characterization of the submicroscopic reservoir in rural Bagamoyo district. A map of Bagamoyo district in eastern Tanzania depicts the size of the submicroscopic reservoir among 15 villages and the locations of the two clinics and two schools where study participants were recruited. The submicroscopic malaria ratio is the ratio of PCR+/RDT-cases to all PCR+ cases. in the village. Zemba village is not included in the map, but is located nearby Mwavi proper village based on the Tanzania Regional Postcode list.

Among these 15 villages in Bagamoyo district, the proportion of malaria-positive persons with subpatent infection, termed the submicroscopic ratio, ranged from 45% to 92% (Table 2). In general, as PCR prevalence increased, the prevalence of malaria detected by microscopy increased in a linear fashion (r^2^ = 0.72, *p* < 0.001) (Figure 3A). Meanwhile, the submicroscopic ratio decreased as overall malaria prevalence (based on PCR) increased (r^2^ = 0.49, *p* = 0.004) (Figure 3B). This finding of a larger submicroscopic reservoir in villages with lower malaria transmission remained statistically significant after adjusting for the number of participants per village, the median age of participants per village, and the proportion of females in each village (*p* = 0.003). There was no difference in the submicroscopic ratio based on whether villages were clustered closer to either of the two main study sites.

**Table 2.**
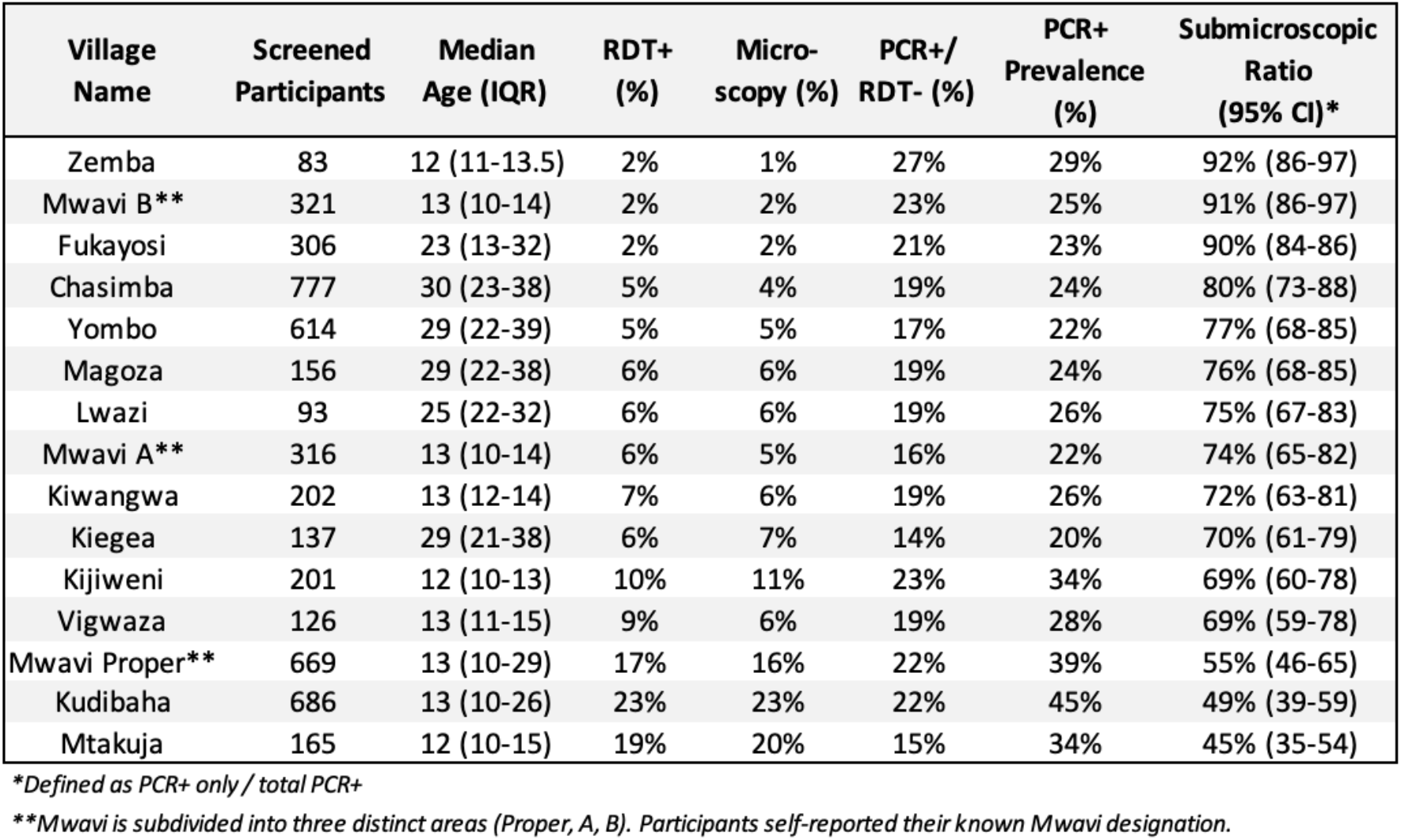
Village-level malaria prevalence among participants recruited from 15 villages in rural Bagamoyo district. Villages are listed in descending order according to the submicroscopic ratio. The proportion of PCR-positive malaria that was RDT-negative, termed the submicroscopic ratio, ranges from 45-92% and is inversely associated with malaria prevalence.

**Figure 3.**
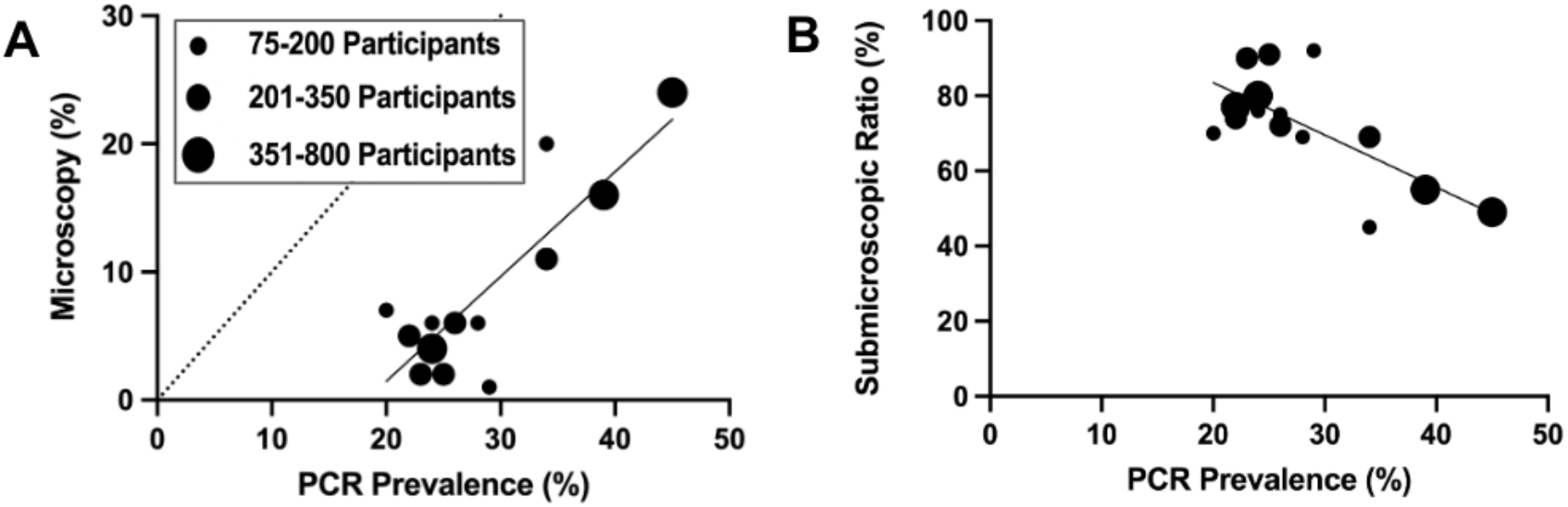
Village-level characterization of malaria prevalence and the size of the submicroscopic malaria. Each data point represents a village, with size corresponding to the number participants reporting residence in that village. Microscopic malaria prevalence from the sampled villages correlated with PCR malaria prevalence (A). The size of the submicroscopic reservoir for each village, indicated by the submicroscopic ratio (PCR+ only cases/total PCR+ cases), increases with decreasing malaria prevalence (B).

### Natural history of submicroscopic parasitemia

From April 2019 to November 2021, 313 participants with submicroscopic malaria were enrolled for weekly monitoring to observe the natural history of submicroscopic parasitemia over 4 weeks. If they developed symptomatic or RDT-positive malaria, they were promptly treated, while those who remained asymptomatic had parasite assessments at weeks 2 and 4 to evaluate the proportion with persistent submicroscopic carriage (Figure 4). Persons in this cohort had an initial median parasite density of 2.8 parasites/μL at enrollment (as estimated by PCR). Among the 295 followed to week 2, 16% (47/295) developed RDT-positive malaria and were treated. Of these 47, roughly half were symptomatic at treatment, with 12/47 (26%) becoming symptomatic before the week 2 study visit and 15/47 (32%) being symptomatic at week 2. The remaining 20/47 (43%) were asymptomatic at the time of treatment. Another 33% (96) of the 295 followed to week 2 had PCR evidence of persistent submicroscopic malaria at week 2, while roughly half (145, 49%) seemingly resolved their parasitemia. However, at least 21% of these (30/145) likely harbored low density parasitemia below the limit of detection of our PCR assay, as they were again PCR-positive at week 4. By week 4, among those not lost to follow-up (LTFU), another 6% (14/219), developed RDT-positive malaria. Cumulatively after 4 weeks, one-quarter of submicroscopic parasite carriers (61/266, 23%) developed patent malaria, one-quarter (67, 25%) continued with submicroscopic asymptomatic carriage, and half (133, 50%) became PCR-negative.

**Figure 4.**
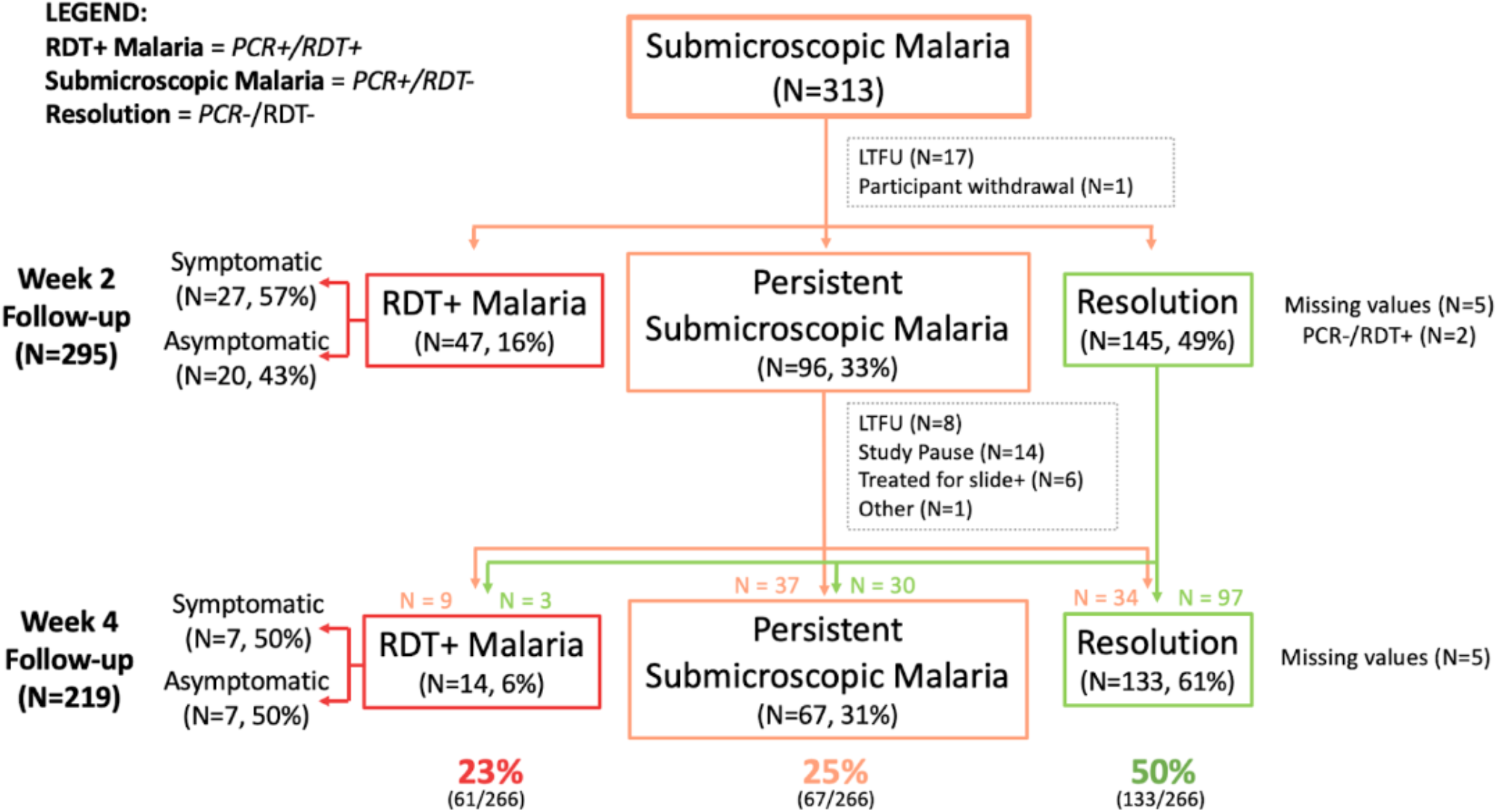
The natural history of submicroscopic parasite carriers over 4 weeks follow-up. The flow diagram depicts the parasite status (based on RDT and PCR) of those who screened positive for submicroscopic malaria (PCR+/RDT-), then followed without treatment and assessed 2 and 4 weeks later. Those who developed RDT+ malaria were treated and discharged. In total, of those enrolled and followed to week 4 (n=219) or became RDT+ before then (n=47) (total n=266), 23% of submicroscopic carriers developed RDT-positive malaria, 25% continued to have asymptomatic submicroscopic carriage, and the remaining had spontaneous resolution by week 4, though this last group may have continued to harbor low density infection not detected by PCR. LFTU indicates lost to follow-up.

As would be expected, a higher initial parasite density was associated with progression to patent malaria within 4 weeks (median 4.5 vs. 2.6 p/μL in those with persistent or resolution of parasitemia at 4 weeks, *p* = 0.03), and the development of patent malaria usually occurred with a rising parasite density (Supplemental Figure 2).

Though malaria-positive school age children (ages 6-17) were overall less likely to have submicroscopic infection compared to adults, those with submicroscopic parasitemia had similar initial parasite densities to the adults in this cohort (median 2.8p/uL vs. 2.7p/uL), and they were not more likely to develop patent malaria compared to adults (28% (31/109) vs. 20% (30/152), *p* = 0.14), though this comparison may be limited by sample size.

Overall, parasitological outcomes at week 4 were not associated with gender, BMI, season (wet vs. dry), reported number of malaria episodes in the past year, or gametocyte carriage at enrollment. However, transmission intensity, as indicated by village-level malaria prevalence, was associated with progression to patent malaria. A higher PCR prevalence in the village in which a participant resided was associated with an increased odds of developing patent malaria. For every 5% percent increase in village-level PCR prevalence, the odds of a submicroscopic carrier developing RDT-positive infection increased by a factor of 1.3 (*p* = 0.006). Of the 110 persons living in high-prevalence villages, 28% developed patent malaria within 4 weeks, compared to 14% of the 119 persons living in medium or low-prevalence villages (Figure 5).

**Figure 5.**
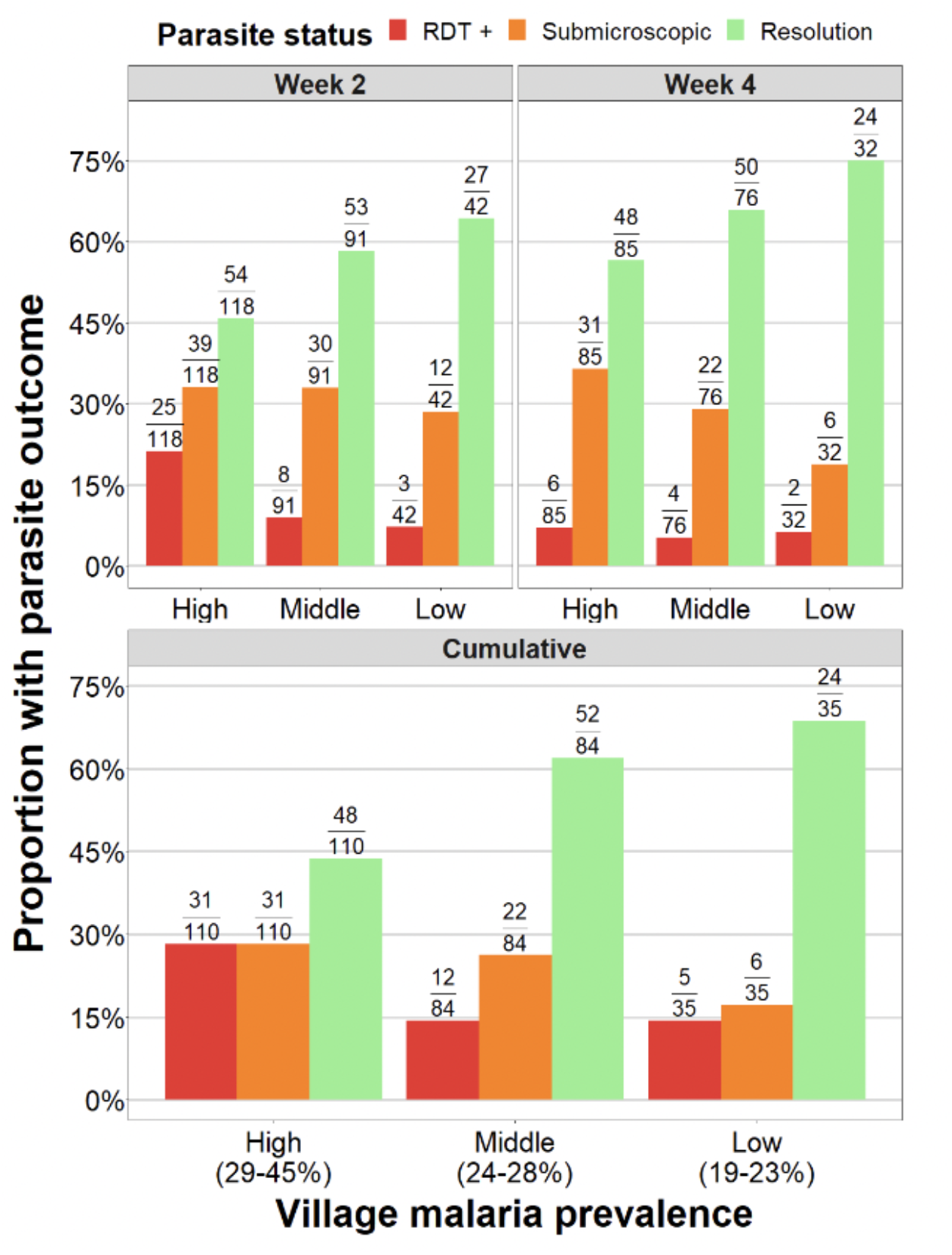
Parasitologic outcomes for untreated submicroscopic parasitemia after 2 and 4 weeks of follow-up, stratified by village malaria prevalence. The proportion of participants with asymptomatic submicroscopic malaria who became RDT-positive (red), continued to have asymptomatic submicroscopic infection (orange), or became PCR-negative (green) is depicted after 2 weeks and 4 weeks of follow-up, with cumulative outcomes depicted in the bottom panel. The minority of participants who developed RDT+ patent malaria were more likely to reside in villages with higher malaria prevalence.

## Discussion

We sought to understand how the submicroscopic reservoir changes as the malaria landscape evolves. In a study employing school and clinic-based cross-sectional screening over three years, we found that in an area of coastal Tanzania where transmission has declined over the last 15 years, roughly 28% of asymptomatic school age children and adults still harbor malaria, with two-thirds of these asymptomatically infected individuals harboring submicroscopic infection. While many studies have highlighted the predominance of asymptomatic low density infections in areas of lower or declining malaria transmission, few have specifically examined factors associated with submicroscopic carriage (vs. RDT-detectable patent malaria), including both individual and ecological factors (Pava et al., 2016; Whittaker et al., 2021). Factors associated with submicroscopic carriage in our study likely relate to acquired immunity (increased age or adult status, an association that is well described) (Doolan et al., 2009; Robortella et al., 2020; Whittaker et al., 2021) and decreased mosquito exposure (bednets and screened windows). Interestingly, females were more likely to be submicroscopic carriers (OR 1.6 (1.3-2.0)) a finding that was also mirrored in Papua, Indonesia, among a population with a similar rate of asymptomatic and submicroscopic infection with a similar odds ratio (adjusted OR = 1.4 (1.0– 1.8)) (Pava et al., 2016). This may be consistent with recent observations that females have higher anti-malaria antibody titers (Chan et al., 2022) and clear asymptomatic infections at a faster rate than males (Briggs et al., 2020).

In a global meta-analysis of the submicroscopic reservoir incorporating 259 surveys, a small but significant increase in the submicroscopic ratio was detected during dry season sampling, with submicroscopic infections less common during the wet season than the dry season (Whittaker et al., 2021). We found a similar association between dry months (based on monthly total rainfall) and submicroscopic carriage, but this association was not significant until we incorporated a 9-week “lag” to account for the time it takes for rainfall to affect mosquito populations and malaria transmission (Bisanzio et al., 2023; Krefis et al., 2011) (Table 1). Finally, submicroscopic carriage appeared to increase over the years of the study, perhaps driven by decreased rainfall but we are unable to rigorously assess this. One might speculate that increased screening and treatment of RDT-positive cases as part of our study may have contributed to increased submicroscopic carriage over time, but i) the discontinuous sampling in early years of the study and a long screening pause before the last year of the study argue against this and ii) the absolute numbers of treated RDT-positive cases was relatively small, especially in the last year of the study (Supplemental Figure 1).

A key novel observation in our study is that village-level prevalence, the spectrum of which reflects the “micro-heterogeneity” of transmission, shapes the size of the submicroscopic reservoir. Overall, 68% of identified parasite carriers had submicroscopic infection, but this proportion differed dramatically based on the village-level PCR prevalence of where participants resided (45-92% in Table 2). Since PCR prevalence correlates well with other measures of transmission intensity (Manjurano et al., 2011), this suggests an association between transmission intensity and the size of the submicroscopic reservoir: as transmission declines, there is a decrease in the total number of parasite carriers, but a larger proportion of these are difficult-to-detect submicroscopic carriers. This has been demonstrated on large regional scales (Whittaker et al., 2021), but to our knowledge, this has not been explicitly studied on a geographically smaller scale (Idris et al., 2016; Robortella et al., 2020; Rosas-Aguirre et al., 2017). Our natural history data, combined with evidence from other recent studies of submicroscopic malaria, may point to why this may be the case. While one might expect individuals living in higher transmission settings to develop more acquired immunity and thus be more likely to control low density parasitemia, they are also more likely to be super-infected with multiple infective bites over time. If the development of patent or symptomatic malaria often arises from superinfection (i.e., a new infected clone or clones) (Sumner et al., 2021a, 2021b), rather than rising density of chronically circulating clones, then it follows that higher transmission intensity would be associated with what appears to be a higher rate of progression to patent malaria and clinical symptoms (Sumner et al., 2021b), as we found. We are currently pursuing genotyping of our natural history cohort to better understand whether patent infections arising over the course of follow-up represented new strains vs. persistent strains responsible for chronic carriage.

The findings presented are subject to multiple limitations. First, sampling was not population-based but was rather convenience sampling at schools and clinics, resulting in recruitment of more females, and excluded children under 6 years of age. Nevertheless, a diverse sample was achieved, and differences in age and gender distribution across villages did not account for differences in the size of submicroscopic reservoir. Second, though the study spanned multiple years, sampling was discontinuous and we were limited in our ability to investigate the effect of time on our variables of interest. Third, while we excluded those with recent antimalarial use, this was by self-report, and an unknown proportion of low-density infections may have resulted from prior partial treatment (Andagalu et al., 2022; Lindblade et al., 2013). Ultimately, it is hard to overcome heterogeneity in the etiology and prior duration or “age” of a submicroscopic infection without a longitudinal study design that allows one to capture incident infection (Andolina et al., 2023; Barry et al., 2021). Fourth, we did not perform molecular gametocyte detection on the large screening cohort to better understand their infectious potential; results of mosquito feeding studies in a subset of PCR-positive participants is forthcoming (“ASTMH-2022-Annual-Meeting-Program-Book_1.pdf,” n.d.). Fifth, in our natural history study we used RDT positivity (patent malaria) regardless of symptoms as a trigger for treatment. Half of those with RDT-positivity at week 2 remained asymptomatic, and it is unclear if they would have remained asymptomatic or later resolved their parasitemia. Finally, parasitological outcomes would be better characterized by molecular genotyping to distinguish persisting and newly infected strains; this work is ongoing.

In conclusion, this work highlights the complex interplay of host and environmental factors shaping mosquito-borne transmission intensity and acquired immunity that makes malaria epidemiology hyper-local, in turn shaping the submicroscopic reservoir. Increasingly, as transmission declines, how best to identify and target pockets or hotspots of at-risk populations remains a key research question (Andolina et al., 2021; Bath et al., 2021; Bousema et al., 2016; Collins et al., 2019; Cotter et al., 2013; Dabira et al., 2022; Hsiang et al., 2020; Sturrock et al., 2013). Studying the submicroscopic reservoir is important both for the clinical implications of subclinical carriage (Chen et al., 2016; Lindblade et al., 2013; van Eijk et al., 2023) as well as the contribution of submicroscopic carriers to the infectious reservoir, i.e., their role in sustaining transmission (Andolina et al., 2023, 2021; Gonçalves et al., 2017; Lin et al., 2014). As more countries progress towards malaria elimination, a better understanding of the factors that shape the submicroscopic reservoir can help guide interventions to maintain progress towards healthy populations free of malaria.

## Data Availability

All data produced in the present study are available upon reasonable request to the authors.

## Acknowledgements

We are grateful to the study participants as well as the staff at the schools and health centers in Bagamoyo for their support. We also thank the study team at Muhimbili University of Health and Allied Sciences (MUHAS) for carrying out the field activities.

## References

Andagalu B, Watson OJ, Onyango I, Opot B, Okoth R, Chemwor G, Sifuna P, Juma D, Cheruiyot A, Yeda R, Okudo C, Wafubwa J, Yalwala S, Abuom D, Ogutu B, Cowden J, Akala HM, Kamau E. 2022. Malaria Transmission Dynamics in High Transmission Setting of Western Kenya and the Inadequate Treatment Response to Artemether-lumefantrine in an Asymptomatic Population. Clin Infect Dis 76:704–712.

Andolina C, Ramjith J, Rek J, Lanke K, Okoth J, Grignard L, Arinaitwe E, Briggs J, Bailey J, Aydemir O, Kamya MR, Greenhouse B, Dorsey G, Staedke SG, Drakeley C, Jonker M, Bousema T. 2023. Plasmodium falciparum gametocyte carriage in longitudinally monitored incident infections is associated with duration of infection and human host factors. Sci Rep 13:7072.

Andolina C, Rek JC, Briggs J, Okoth J, Musiime A, Ramjith J, Teyssier N, Conrad M, Nankabirwa JI, Lanke K, Rodriguez-Barraquer I, Meerstein-Kessel L, Arinaitwe E, Olwoch P, Rosenthal PJ, Kamya MR, Dorsey G, Greenhouse B, Drakeley C, Staedke SG, Bousema T. 2021. Sources of persistent malaria transmission in a setting with effective malaria control in eastern Uganda: a longitudinal, observational cohort study. Lancet Infect Dis 21:1568–1578.

ASTMH-2022-Annual-Meeting-Program-Book_1.pdf. n.d.

Barry A, Bradley J, Stone W, Guelbeogo MW, Lanke K, Ouedraogo A, Soulama I, Nébié I, Serme SS, Grignard L, Patterson C, Wu L, Briggs JJ, Janson O, Awandu SS, Ouedraogo M, Tarama CW, Kargougou D, Zongo S, Sirima SB, Marti M, Drakeley C, Tiono AB, Bousema T. 2021. Higher gametocyte production and mosquito infectivity in chronic compared to incident Plasmodium falciparum infections. Nat Commun 12:2443.

Bath D, Cook J, Govere J, Mathebula P, Morris N, Hlongwana K, Raman J, Seocharan I, Zitha A, Zitha M, Mabuza A, Mbokazi F, Machaba E, Mabunda E, Jamesboy E, Biggs J, Drakeley C, Moonasar D, Maharaj R, Coetzee M, Pitt C, Kleinschmidt I. 2021. Effectiveness and cost-effectiveness of reactive, targeted indoor residual spraying for malaria control in low-transmission settings: a cluster-randomised, noninferiority trial in South Africa. Lancet 397:816–827.

Berzosa P, de Lucio A, Romay-Barja M, Herrador Z, González V, García L, Fernández-Martínez A, Santana-Morales M, Ncogo P, Valladares B, Riloha M, Benito A. 2018. Comparison of three diagnostic methods (microscopy, RDT, and PCR) for the detection of malaria parasites in representative samples from Equatorial Guinea. Malar J 17:333.

Bhatt S, Weiss DJ, Cameron E, Bisanzio D, Mappin B, Dalrymple U, Battle K, Moyes CL, Henry A, Eckhoff PA, Wenger EA, Briët O, Penny MA, Smith TA, Bennett A, Yukich J, Eisele TP, Griffin JT, Fergus CA, Lynch M, Lindgren F, Cohen JM, Murray CLJ, Smith DL, Hay SI, Cibulskis RE, Gething PW. 2015. The effect of malaria control on Plasmodium falciparum in Africa between 2000 and 2015. Nature 526:207–211.

Bisanzio D, Lalji S, Abbas FB, Ali MH, Hassan W, Mkali HR, Al-Mafazy A-W, Joseph JJ, Nyinondi S, Kitojo C, Serbantez N, Reaves E, Eckert E, Ngondi JM, Reithinger R. 2023. Spatiotemporal dynamics of malaria in Zanzibar, 2015-2020. BMJ Glob Health 8:e009566.

Bousema T, Stresman G, Baidjoe AY, Bradley J, Knight P, Stone W, Osoti V, Makori E, Owaga C, Odongo W, China P, Shagari S, Doumbo OK, Sauerwein RW, Kariuki S, Drakeley C, Stevenson J, Cox J. 2016. The impact of hotspot-targeted interventions on malaria transmission in Rachuonyo South District in the western Kenyan highlands: A cluster-randomized controlled trial. PLoS Med 13:e1001993.

Briggs J, Teyssier N, Nankabirwa JI, Rek J, Jagannathan P, Arinaitwe E, Bousema T, Drakeley C, Murray M, Crawford E, Hathaway N, Staedke SG, Smith D, Rosenthal PJ, Kamya M, Dorsey G, Rodriguez-Barraquer I, Greenhouse B. 2020. Sex-based differences in clearance of chronic Plasmodium falciparum infection. Elife 9. doi:10.7554/eLife.59872.

Chan J-A, Loughland JR, de la Parte L, Okano S, Ssewanyana I, Nalubega M, Nankya F, Musinguzi K, Rek J, Arinaitwe E, Tipping P, Bourke P, Andrew D, Dooley N, SheelaNair A, Wines BD, Hogarth PM, Beeson JG, Greenhouse B, Dorsey G, Kamya M, Hartel G, Minigo G, Feeney M, Jagannathan P, Boyle MJ. 2022. Age-dependent changes in circulating Tfh cells influence development of functional malaria antibodies in children. Nat Commun 13:4159.

Chen I, Clarke SE, Gosling R, Hamainza B, Killeen G, Magill A, O’Meara W, Price RN, Riley EM. 2016. “Asymptomatic” Malaria: A Chronic and Debilitating Infection That Should Be Treated. PLoS Med 13:e1001942.

Collins KA, Ouedraogo A, Guelbeogo WM, Awandu SS, Stone W, Soulama I, Ouattara MS, Nombre A, Diarra A, Bradley J, Selvaraj P, Gerardin J, Drakeley C, Bousema T, Tiono A. 2019. Investigating the impact of enhanced community case management and monthly screening and treatment on the transmissibility of malaria infections in Burkina Faso: study protocol for a cluster-randomised trial. BMJ Open 9:e030598.

Cotter C, Sturrock HJW, Hsiang MS, Liu J, Phillips AA, Hwang J, Gueye CS, Fullman N, Gosling RD, Feachem RGA. 2013. The changing epidemiology of malaria elimination: new strategies for new challenges. Lancet 382:900–911.

Dabira ED, Soumare HM, Conteh B, Ceesay F, Ndiath MO, Bradley J, Mohammed N, Kandeh B, Smit MR, Slater H, Peeters Grietens K, Broekhuizen H, Bousema T, Drakeley C, Lindsay SW, Achan J, D’Alessandro U. 2022. Mass drug administration of ivermectin and dihydroartemisinin-piperaquine against malaria in settings with high coverage of standard control interventions: a cluster-randomised controlled trial in The Gambia. Lancet Infect Dis 22:519–228.

Das S, Jang IK, Barney B, Peck R, Rek JC, Arinaitwe E, Adrama H, Murphy M, Imwong M, Ling CL, Proux S, Haohankhunnatham W, Rist M, Seilie AM, Hanron A, Daza G, Chang M, Nakamura T, Kalnoky M, Labarre P, Murphy SC, McCarthy JS, Nosten F, Greenhouse B, Allauzen S, Domingo GJ. 2017. Performance of a High-Sensitivity Rapid Diagnostic Test for Plasmodium falciparum Malaria in Asymptomatic Individuals from Uganda and Myanmar and Naive Human Challenge Infections. Am J Trop Med Hyg 97:1540–1550.

Doolan DL, Dobaño C, Baird JK. 2009. Acquired immunity to malaria. Clin Microbiol Rev 22:13–36.

Gamoyo M, Reason C, Obura D. 2015. Rainfall variability over the East African coast. Theor Appl Climatol 120:1–2.

Global messaging: World malaria report 2022. n.d. https://www.who.int/publications/m/item/WHO-UCN-GMP-2022.07.

Gonçalves BP, Kapulu MC, Sawa P, Guelbéogo WM, Tiono AB, Grignard L, Stone W, Hellewell J, Lanke K, Bastiaens GJH, Bradley J, Nébié I, Ngoi JM, Oriango R, Mkabili D, Nyaurah M, Midega J, Wirth DF, Marsh K, Churcher TS, Bejon P, Sirima SB, Drakeley C, Bousema T. 2017. Examining the human infectious reservoir for Plasmodium falciparum malaria in areas of differing transmission intensity. Nat Commun 8:1133.

Hsiang MS, Ntuku H, Roberts KW, Dufour M-SK, Whittemore B, Tambo M, McCreesh P, Medzihradsky OF, Prach LM, Siloka G, Siame N, Gueye CS, Schrubbe L, Wu L, Scott V, Tessema S, Greenhouse B, Erlank E, Koekemoer LL, Sturrock HJW, Mwilima A, Katokele S, Uusiku P, Bennett A, Smith JL, Kleinschmidt I, Mumbengegwi D, Gosling R. 2020. Effectiveness of reactive focal mass drug administration and reactive focal vector control to reduce malaria transmission in the low malaria-endemic setting of Namibia: a cluster-randomised controlled, open-label, two-by-two factorial design trial. Lancet 395:1361–1373.

Idris ZM, Chan CW, Kongere J, Gitaka J, Logedi J, Omar A, Obonyo C, Machini BK, Isozumi R, Teramoto I, Kimura M, Kaneko A. 2016. High and Heterogeneous Prevalence of Asymptomatic and Sub-microscopic Malaria Infections on Islands in Lake Victoria, Kenya. Sci Rep 6:36958.

Japan International Cooperation Agency (JICA), Global Environment Department. 2005. The Study on Water Supply Improvement in Coast Region and Dar es Salaam Peri-Urban in the United Republic of Tanzania: Final Report Data Book. JICA.

Krefis AC, Schwarz NG, Krüger A, Fobil J, Nkrumah B, Acquah S, Loag W, Sarpong N, Adu-Sarkodie Y, Ranft U, May J. 2011. Modeling the relationship between precipitation and malaria incidence in children from a holoendemic area in Ghana. Am J Trop Med Hyg 84:285–291.

Leski TA, Taitt CR, Swaray AG, Bangura U, Reynolds ND, Holtz A, Yasuda C, Lahai J, Lamin JM, Baio V, Jacobsen KH, Ansumana R, Stenger DA. 2020. Use of real-time multiplex PCR, malaria rapid diagnostic test and microscopy to investigate the prevalence of Plasmodium species among febrile hospital patients in Sierra Leone. Malar J 19:84.

Lindblade KA, Steinhardt L, Samuels A, Kachur SP, Slutsker L. 2013. The silent threat: asymptomatic parasitemia and malaria transmission. Expert Rev Anti Infect Ther 11:623–639.

Lin JT, Saunders DL, Meshnick SR. 2014. The role of submicroscopic parasitemia in malaria transmission: what is the evidence? Trends Parasitol 30:183–190.

Maina J, Ouma PO, Macharia PM, Alegana VA, Mitto B, Fall IS, Noor AM, Snow RW, Okiro EA. 2019. A spatial database of health facilities managed by the public health sector in sub Saharan Africa. Sci Data 6:134.

Manjurano A, Okell L, Lukindo T, Reyburn H, Olomi R, Roper C, Clark TG, Joseph S, Riley EM, Drakeley C. 2011. Association of sub-microscopic malaria parasite carriage with transmission intensity in north-eastern Tanzania. Malar J 10:370.

Markwalter CF, Ngasala B, Mowatt T, Basham C, Park Z, Loya M, Muller M, Plowe C, Nyunt M, Lin JT. 2021. Direct Comparison of Standard and Ultrasensitive PCR for the Detection of Plasmodium falciparum from Dried Blood Spots in Bagamoyo, Tanzania. Am J Trop Med Hyg 104:1371–1374.

Mfuh KO, Achonduh-Atijegbe OA, Bekindaka ON, Esemu LF, Mbakop CD, Gandhi K, Leke RGF, Taylor DW, Nerurkar VR. 2019. A comparison of thick-film microscopy, rapid diagnostic test, and polymerase chain reaction for accurate diagnosis of Plasmodium falciparum malaria. Malar J 18:73.

Ministry of Health, The United Republic of Tanzania. 2021. Tanzania Health Facility Registry (HFR). Mwanzo. n.d. https://www.meteo.go.tz/.

Okell LC, Bousema T, Griffin JT, Ouédraogo AL, Ghani AC, Drakeley CJ. 2012. Factors determining the occurrence of submicroscopic malaria infections and their relevance for control. Nat Commun 3:1237.

OpenStreetMap contributors, Geofabrik GmbH. 2021. Tanzania OpenStreetMap data. OpenStreetMap. https://download.geofabrik.de/africa/tanzania.html.

Pava Z, Burdam FH, Handayuni I, Trianty L, Utami RAS, Tirta YK, Kenangalem E, Lampah D, Kusuma A, Wirjanata G, Kho S, Simpson JA, Auburn S, Douglas NM, Noviyanti R, Anstey NM, Poespoprodjo JR, Marfurt J, Price RN. 2016. Submicroscopic and Asymptomatic Plasmodium Parasitaemia Associated with Significant Risk of Anaemia in Papua, Indonesia. PLoS One 11:e0165340.

Robortella DR, Calvet AA, Amaral LC, Fantin RF, Guimarães LFF, França Dias MH, Brito CFA de, Sousa TN de, Herzog MM, Oliveira-Ferreira J, Carvalho LH. 2020. Prospective assessment of malaria infection in a semi-isolated Amazonian indigenous Yanomami community: Transmission heterogeneity and predominance of submicroscopic infection. PLoS One 15:e0230643.

Rosas-Aguirre A, Guzman-Guzman M, Gamboa D, Chuquiyauri R, Ramirez R, Manrique P, Carrasco-Escobar G, Puemape C, Llanos-Cuentas A, Vinetz JM. 2017. Micro-heterogeneity of malaria transmission in the Peruvian Amazon: a baseline assessment underlying a population-based cohort study. Malar J 16:312.

Rutta ASM, Francis F, Mmbando BP, Ishengoma DS, Sembuche SH, Malecela EK, Sadi JY, Kamugisha ML, Lemnge MM. 2012. Using community-owned resource persons to provide early diagnosis and treatment and estimate malaria burden at community level in north-eastern Tanzania. Malar J 11:152.

Sturrock HJW, Hsiang MS, Cohen JM, Smith DL, Greenhouse B, Bousema T, Gosling RD. 2013. Targeting asymptomatic malaria infections: active surveillance in control and elimination. PLoS Med 10:e1001467.

Sumner KM, Freedman E, Mangeni JN, Obala AA, Abel L, Edwards JK, Emch M, Meshnick SR, Pence BW, Prudhomme-O’Meara W, Taylor SM. 2021a. Exposure to Diverse Plasmodium falciparum Genotypes Shapes the Risk of Symptomatic Malaria in Incident and Persistent Infections: A Longitudinal Molecular Epidemiologic Study in Kenya. Clin Infect Dis 73:1176–1184.

Sumner KM, Mangeni JN, Obala AA, Freedman E, Abel L, Meshnick SR, Edwards JK, Pence BW, Prudhomme-O’Meara W, Taylor SM. 2021b. Impact of asymptomatic Plasmodium falciparum infection on the risk of subsequent symptomatic malaria in a longitudinal cohort in Kenya. elife 10.

Tanzania Communications Regulatory Authority (TCRA). 2019. Tanzania Postal Codes-Tanzania Zip codes. Udahiliportal.com.

Tarimo BB, Nyasembe VO, Ngasala B, Basham C, Rutagi IJ, Muller M, Chhetri SB, Rubinstein R, Juliano JJ, Loya M, Dinglasan RR, Lin JT, Mathias DK. 2022. Seasonality and transmissibility of Plasmodium ovale in Bagamoyo District, Tanzania. Parasit Vectors 15:56.

van Eijk AM, Stepniewska K, Hill J, Taylor SM, Rogerson SJ, Cottrell G, Chico RM, Gutman JR, Tinto H, Unger HW, Yanow SK, Meshnick SR, Ter Kuile FO, Mayor A, Subpatent Malaria in Pregnancy Group. 2023. Prevalence of and risk factors for microscopic and submicroscopic malaria infections in pregnancy: a systematic review and meta-analysis. Lancet Glob Health 11:e1061–e1074.

Whittaker C, Slater H, Nash R, Bousema T, Drakeley C, Ghani AC, Okell LC. 2021. Global patterns of submicroscopic Plasmodium falciparum malaria infection: insights from a systematic review and metaanalysis of population surveys. Lancet Microbe 2:e366–e374.

