## Supplemental Figures for "Micro-heterogeneity of transmission shapes the submicroscopic malaria reservoir in coastal Tanzania"

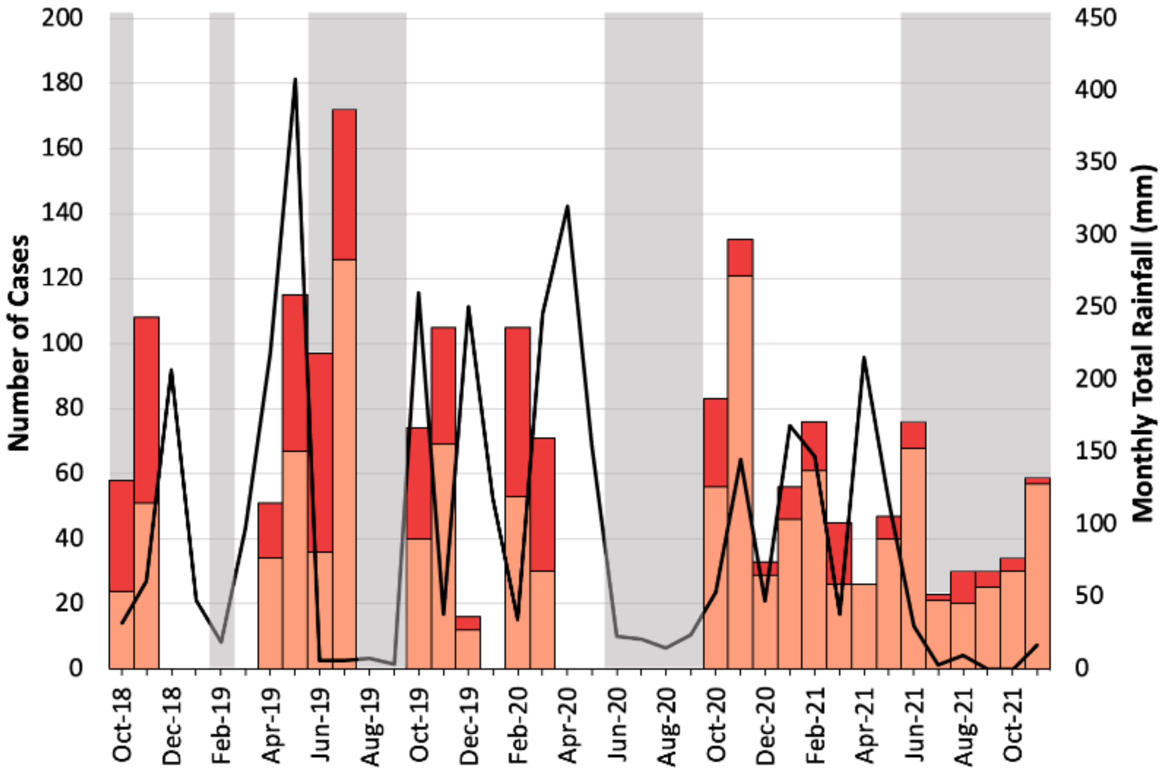


**Supplementary Figure 1**. Number of RDT-positive and submicroscopic malaria cases identified during screening. Data are the same as in Figure 1 but show the actual numbers of cases that screened positive over time.

**Supplementary Table 1. Participant characteristics**

| | n | % |
|---|---|---|
| ***SCREENED PARTICIPANTS*** | 6076 | - |
| **Age (Median, IQR)** | 19 (12-31) | - |
| **Gender** | | |
| Male | 1904 | 31% |
| Female | 4171 | 69% |
| **RDT+** | 554 | 9% |
| **Microscopy+** | 525 | 9% |
| **PCR+/RDT-** | 1168 | 19% |
| **PCR+** | 1722 | 28% |
| ***ENROLLED PARTICIPANTS*** | 544 | 9% |
| **Age (Median, IQR)** | 18 (12-31) | - |
| **Gender** | | |
| Male | 197 | 36% |
| Female | 347 | 64% |
| **RDT+** | 217 | 40% |
| **Microscopy+** | 188 | 35% |
| **PCR+/RDT-** | 327 | 60% |
| **PCR+** | 544 | 100% |
| **Mosquito net use last night (n = 532)** | 483 | 89% |
| **Water within 2 minute walk (n = 530)** | 62 | 11% |
| **Roof material** | | |
| Thatching/leaves | 48 | 9% |
| Palm leaves/bamboo | 12 | 2% |
| Sheet metal | 479 | 88% |
| **Housing material** | | |
| Earth | 33 | 6% |
| Bamboo with mud | 369 | 68% |
| Stones with mud | 3 | 1% |
| Cement | 123 | 23% |
| **Windows** | | |
| Screens | 197 | 36% |
| Open | 235 | 43% |
| Planks | 10 | 2% |
| Other* | 103 | 19% |

**Other = none, glass, or unknown*

**Supplementary Table 2. Participant Characteristics Stratified by Study Time Period**

| | 2A. October 2018 - March 2020 | | | | | 2B. October 2020 - November 2021 | | | | |
|---|---|---|---|---|---|---|---|---|---|---|
| | Total (n) | PCR+/RDT- (n, %) | PCR+/RDT+ (n, %) | OR (95% CI) | *p* value | Total (n) | PCR+/RDT- (n, %) | PCR+/RDT+ (n, %) | OR (95% CI) | *p* value |
| **Age (Median, IQR)** | | 21 (12-32) | 13 (10-20) | | <0.001 | | 21 (12-32) | 16 (13-27) | | 0.2 |
| **Adult status** | | | | | | | | | | |
| ≥18 y.o. | 446 | 309 | 137 | 2.8 (2.2-3.7) | <0.001 | 402 | 343 | 59 | 1.3 (0.9-2.0) | 0.1 |
| <18 y.o. | 526 | 233 | 293 | 1.0 | | 348 | 283 | 65 | 1.0 | |
| **Sex** | | | | | | | | | | |
| Female | 585 | 351 | 234 | 1.5 (1.2-2.0) | 0.001 | 511 | 435 | 76 | 1.4 (1.0-2.1) | 0.07 |
| ≥18 y.o. | 319 | 226 | 93 | 1.3 (0.8-2.0) | 0.3 | 319 | 274 | 45 | 1.2 (0.6-2.4) | 0.5 |
| <18 y.o. | 266 | 125 | 141 | 1.3 (0.9-1.8) | 0.2 | 192 | 161 | 31 | 1.5 (0.8-2.5) | 0.2 |
| Male | 387 | 191 | 196 | 1.0 | | 239 | 191 | 48 | 1.0 | |
| ≥18 y.o. | 127 | 83 | 44 | 1.0 | | 83 | 69 | 14 | 1.0 | |
| <18 y.o. | 260 | 108 | 152 | 1.0 | | 156 | 122 | 34 | 1.0 | |
| **Number of malaria infections in the past one year (Median, IQR)** | | 1 (0-2) | 2 (1-3) | | 0.007 | | 1 (0-2) | 2 (1-2) | | 0.3 |
| **Seasonality (Current Rainfall)** | | | | | | | | | | |
| Dry | 327 | 186 | 141 | 1.1 (0.8-1.4) | 0.6 | 252 | 221 | 31 | 1.6 (1.0-2.5) | 0.03 |
| Wet | 645 | 356 | 289 | 1.0 | | 498 | 405 | 93 | 1.0 | |
| **Seasonality (Prior Rainfall)** | | | | | | | | | | |
| Dry | 228 | 145 | 83 | 1.4 (1.0-1.9) | 0.04 | 376 | 314 | 62 | 1.0 (0.7-1.5) | 1.0 |
| Wet | 578 | 322 | 256 | 1.0 | | 374 | 312 | 62 | 1.0 | |
| **Windows** | | | | | | | | | | |
| Screen | 99 | 53 | 46 | 1.7 (1.0-2.7) | 0.06 | 98 | 80 | 18 | 1.4 (0.7-2.9) | 0.3 |
| Open | 153 | 63 | 90 | 1.0 | | 82 | 62 | 20 | 1.0 | |
| **Housing** | | | | | | | | | | |
| Bamboo | 225 | 117 | 108 | 1.2 (0.7-2.1) | 0.5 | 144 | 110 | 34 | 0.8 (0.4-1.7) | 0.6 |
| Cement | 72 | 34 | 38 | 1.0 | | 51 | 41 | 10 | 1.0 | |
| **Roof** | | | | | | | | | | |
| Sheet metal | 303 | 152 | 151 | 0.8 (0.4-1.6) | 0.6 | 25 | 15 | 10 | 0.4 (0.2-1.0) | 0.04 |
| Other | 40 | 22 | 18 | 1.0 | | 176 | 138 | 38 | 1.0 | |
| **Mosquito net use last night** | | | | | | | | | | |
| Yes | 291 | 152 | 139 | 1.4 (0.8-2.6) | 0.3 | 192 | 149 | 43 | 4.3 (1.1-17) | <0.05 |
| No | 50 | 22 | 28 | 1.0 | | 9 | 4 | 5 | 1.0 | |
| **Water within 2 minute walk** | | | | | | | | | | |
| Yes | 56 | 28 | 28 | 1.0 (0.6-1.7) | 0.9 | 194 | 148 | 46 | 0.6 (0.0-5.6) | 1.0 |
| No | 284 | 145 | 139 | 1.0 | | 6 | 5 | 1 | 1.0 | |

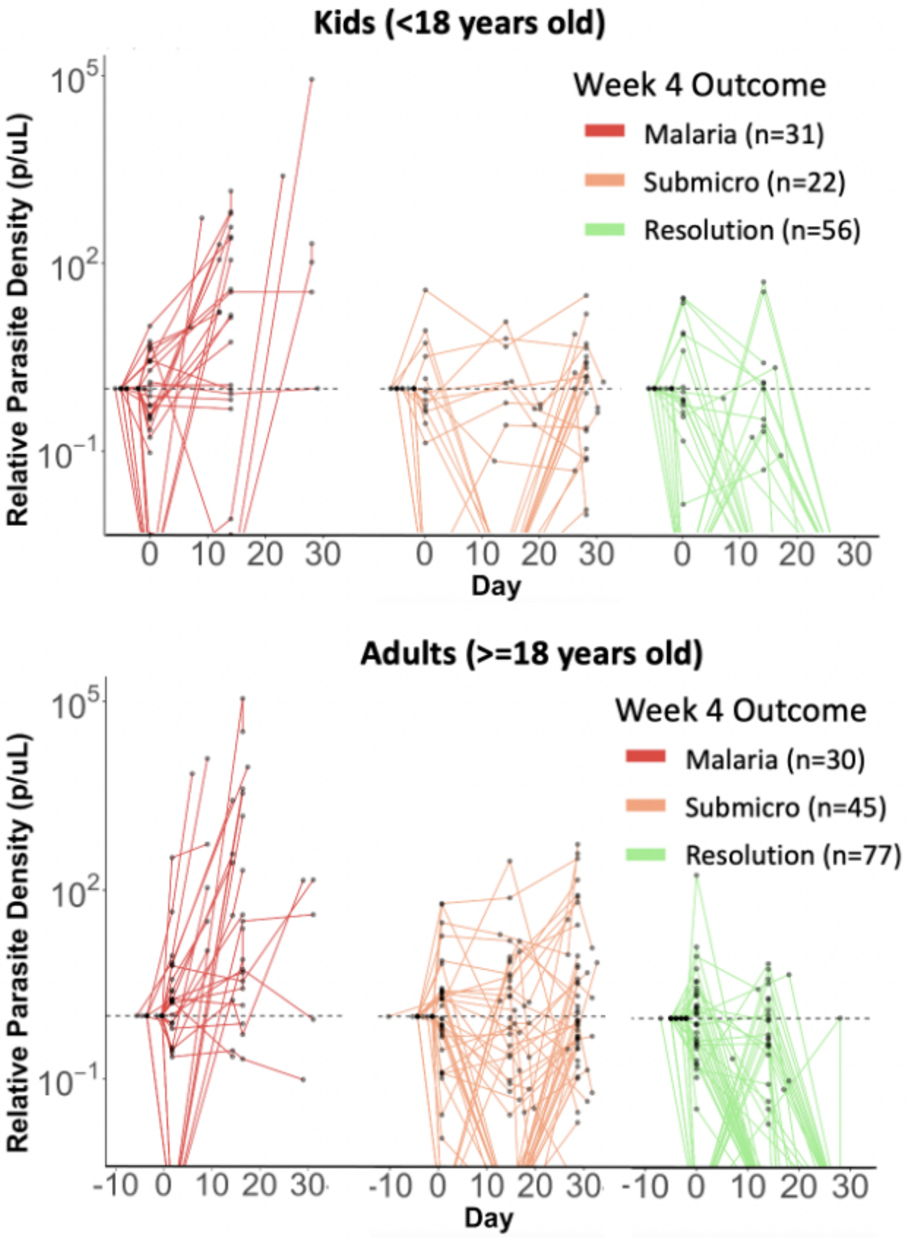


**Supplemental Figure 2. Longitudinal parasite densities among submicroscopic parasite carriers**. Parasite densities relative to screening density (log-10 scale) are depicted among those followed to four weeks before treatment, with treatment sooner if they developed RDT-positive or symptomatic malaria. Within each panel, participants are grouped according to their parasitological outcome at week 4, as outlined in Figure 5. Horizontal dashed line represents y=1
